# Evaluation of an AI-Powered Patient Education Application in Bariatric Surgery: A Prospective Mixed-Methods Feasibility Study

**DOI:** 10.64898/2026.09.16.26363276

**Authors:** Jamil S. Samaan, Stephanie T. Nguyen, Sevag Hamamah, Ashley Tran, Rachel Carmen Ceasar, Nicole Wolfe, Trent Yu, Diana Chavez, Nithya Rajeev, Rashmi Advani, Rabindra Watson, Barham Abu Dayyeh, Kulmeet Sandhu, Harry J. Wong, Nicholas P. Tatonetti, Kamran Samakar

## Abstract

**Background:** Patient education is critical to safe and effective postoperative bariatric surgery care. Large language models (LLMs) may improve access to personalized patient education but evidence regarding direct patient use in clinical settings remains limited. We evaluated BariBot, an LLM-powered educational application for patients undergoing bariatric surgery.

**Methods:** We conducted a prospective, single-arm mixed-methods feasibility study at a tertiary academic medical center from May 2024 to May 2025. Adult patients undergoing bariatric surgery used BariBot independently on postoperative day 1. The application provided conversational postoperative education and adapted language complexity to participants’ educational attainment. Outcomes included feasibility, usability, acceptability, perceived trustworthiness, and anticipated future use. Usability was assessed using the System Usability Scale (SUS). Immediately after BariBot use, participants completed a semi-structured qualitative interview assessing acceptability, perceived trustworthiness, and anticipated future use. Interview transcripts underwent thematic analysis.

**Results:** Sixteen participants completed the BariBot use session, SUS assessment, and post-intervention interview. No technical failures prevented application use, and no participant requested early discontinuation. The median SUS score was 91.3 (IQR, 81.3-98.1), consistent with excellent perceived usability. Qualitative analysis showed that participants valued the application’s immediacy, structured responses, conversational context, ability to address follow-up questions, judgement-free nature, and future availability between clinical encounters. Trust was strengthened by alignment with prior clinical guidance and perceived institutional association, although participants emphasized the need for transparency, clinician oversight, and use as an adjunct rather than a replacement of clinician guidance.

**Conclusions:** BariBot was feasible to implement and was associated with high perceived usability and acceptability postoperatively after bariatric surgery. Larger, longitudinal, and comparative studies are needed to evaluate safety during unsupervised use and determine whether LLM-based educational applications improve patient understanding, engagement, and clinical outcomes.

## INTRODUCTION

Obesity is a major public health challenge, affecting approximately 16% of adults globally and nearly 40% of adults in the United States (1–3) and is associated with cardiometabolic disease, malignancy, and premature mortality. (4–6) Metabolic and bariatric surgery (MBS) is a safe and effective treatment for obesity that produces durable weight loss, resolves obesity-related comorbidities, and improves long-term survival. (7,8) The benefits and safety of MBS depend not only on procedural success but also on patients’ ability to adopt and maintain behavioral, nutritional, and medical regimens, making patient education a core component of bariatric care. (9–11)

Low health literacy has been linked to poorer adherence to surgical postoperative instructions and outcomes, while in bariatric cohorts, it has been associated with higher emergency room visits and reduced weight loss. (12–16) Despite the availability of traditional resources such as handouts, group classes, and web-based education resources, these approaches are often generic, inconsistently updated, and insufficiently personalized. (17–20) These limitations are particularly consequential postoperatively, when questions regarding diet, symptoms, medications, and activity may arise between clinical encounters, when access to the clinical team may be limited. (21–28) As a result, digital platforms and social media are increasingly used by patients to supplement clinical advice, yet information from these sources may be fragmented, inaccurate, or influenced by commercial benefits. (29–32) This reliance on digital content sources underscores the need for innovative strategies to deliver accurate, accessible, trustworthy, and patient-centered education.

Large language models (LLMs) can generate conversational responses tailored to a user’s question and requested level of complexity and may therefore expand access to patient education. (33) Studies have found that foundation models can provide generally accurate and understandable responses to bariatric surgery questions, although these evaluations have relied on simulated inputs or expert review rather than direct patient input. (34–38) LLMs can also generate inaccurate or fabricated information, provide variable responses across prompts, and offer limited transparency regarding the provenance of their outputs. (39–41) Consequently, benchmark accuracy alone cannot establish whether patient-facing applications are usable, acceptable, or clinically appropriate in practice. The feasibility, safety, and perceived value of LLMs as patient-facing tools remain largely untested, particularly in MBS, where educational needs are essential for optimal outcomes. To address this gap, we developed and evaluated BariBot, an AI-powered educational application designed for patients undergoing bariatric surgery.

## METHODS

### Study Design, Setting, and Participants

We conducted a prospective, single-arm mixed-methods feasibility study at a tertiary academic medical center from May 2024 to May 2025. Participants aged 18 years or older scheduled to undergo elective laparoscopic sleeve gastrectomy or Roux-en-Y gastric bypass for weight loss were eligible for inclusion. Patients were excluded if they lacked capacity to provide informed consent, were legally blind, or primarily used a non-English language for communication, as these factors could impair meaningful interaction with the text-based, English-language application.

Patients scheduled for bariatric surgery were identified preoperatively through the institutional electronic medical record. One to two days prior to surgery, a study team member contacted each patient by telephone using a HIPAA-compliant Doximity platform to explain the study purpose and protocol. Participants provided informed consent before completing any study procedures and received no compensation. The study was approved by the Institutional Review Board.

### BariBot Application

BariBot is an LLM-powered application developed using the custom GPT OpenAI platform (OpenAI). (42) The application facilitates patient-facing conversational guidance across a broad range of topics, including but not limited to recovery, diet, medications, physical activity, and postoperative expectations (**Figure 1**). The application was configured to vary the complexity of its language according to each participant’s self-reported highest educational level completed. This feature was intended to address evidence that unmodified LLM responses may exceed recommended readability levels for patient education. (35,36,43,44) Before participant use, 3 study investigators (K.S., J.S.S., and A.T.) tested the application to refine response clarity, clinical appropriateness, and safety. BariBot was not provided with identifying participant information.

**Figure 1:**
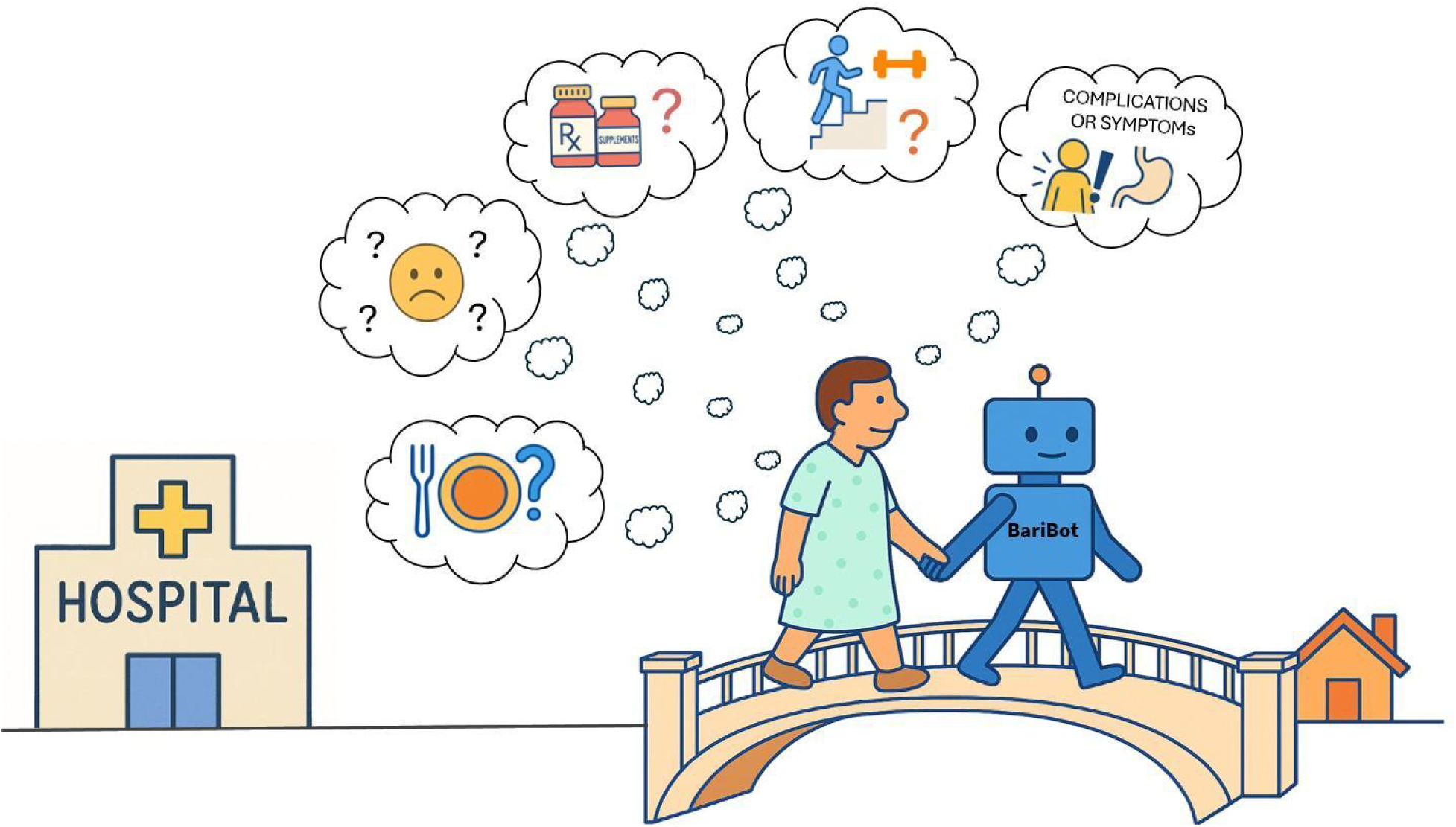
Conceptual illustration of BariBot as a bridge between hospital and home, supporting participants with postoperative questions related to diet, medications, exercise, emotional concerns, and other bariatric surgery-related topics.

### Study Procedures

Patients completed a brief pre-intervention survey assessing demographics and prior experience with artificial intelligence (**Appendix 1: Pre-intervention Survey**). On postoperative day 1 (POD1), participants received an iPad preloaded with the BariBot application. This time point provided a standardized evaluation after immediate postoperative recovery and before hospital discharge.

After a brief technical orientation, participants were encouraged to interact with BariBot independently and had access to the application for up to 5 hours. Study staff were available to address technical problems but did not suggest questions or otherwise direct the content of the conversations. Immediately after the use period, participants completed the System Usability Scale (SUS) and participated in a semi-structured interview.

### Outcomes

The primary objectives were to evaluate the feasibility, usability, acceptability, perceived trustworthiness, and patient anticipated future use of BariBot as a bariatric surgery educational resource. Feasibility was assessed by completion of the BariBot use session, completion of post-intervention assessments, absence of technical failure preventing application use, and absence of participant-requested early discontinuation. Usability was assessed using the SUS, a validated 10-item instrument that generates a composite score ranging from 0 to 100, with higher scores indicating greater perceived usability. (45,46)

Acceptability, perceived trustworthiness, and anticipated future use were evaluated through semi-structured qualitative interviews and thematic analysis. Complete participant– BariBot interaction transcripts were reviewed by a physician for clinical appropriateness.

### Qualitative Data Collection and Analysis

A semi-structured qualitative interview was conducted on postoperative day 1 immediately after BariBot use and completion of the SUS. The interview guide was adapted from a prior study of a healthcare AI application and modified for the MBS context. (**Supplementary Appendix**). (47) The interview guide was designed to explore three overarching domains. To contextualize patient perceptions of BariBot, patients described their experiences and perceptions of educational resources used prior to BariBot. Next, we evaluated patient experience with the application itself, including impressions of BariBot’s accuracy, usefulness, and perceived trustworthiness. Finally, patients reflected on how they envisioned future integration of BariBot and similar AI tools into their postoperative care, including perceptions of broader applicability to other bariatric surgery patients.

Interview transcripts were analyzed using thematic analysis. Three study personnel (SN, JSS, SH) independently reviewed the first three transcripts and developed an initial coding framework through iterative discussion. The framework was refined in consultation with experts in qualitative research (NW, RCC), then applied to subsequent transcripts. A hybrid coding approach was used, combining deductive domains derived from the interview guide with inductively generated codes identified during transcript review. Earlier transcripts were revisited and recoded when necessary to maintain consistency across the dataset. Discrepancies in coding were resolved by consensus. Thematic saturation was defined as the point at which no new concepts or themes emerged in successive interviews and was assessed during iterative transcript review. Coding and data organization were conducted using Microsoft Excel. Representative quotations were selected to illustrate each domain.

### Quantitative Analysis

Quantitative data were analyzed descriptively. Categorical variables are reported as frequencies and percentages. Continuous variables are reported as medians ± interquartile range (IQR) when appropriate. Descriptive analyses were used to summarize participant demographics, pre-intervention information sources, and SUS scores.

## RESULTS

### Feasibility and Usability

All 16 participants completed the pre-intervention survey, BariBot session, SUS assessment, and post-intervention interview. Participant characteristics are shown in **Table 1**. BariBot participant transcripts contained a median of 1,994 words (IQR, 894-4982), and participants submitted a median of 11 inputs per session (IQR, 6.25-22). The most discussed topics were diet (10 participants, 62.5%), pain (9 participants, 56.3%), physical activity or exercise (8 participants, 50%), and recovery or healing (6 participants, 37.5%). No participant requested early discontinuation, and no technical failures prevented application use. The median SUS score was 91.3 (IQR, 81.3-98.1). This exceeded the conventional benchmark for above-average usability and was consistent with excellent perceived usability. (45,46) Physician review of complete participant-BariBot interaction transcripts identified no responses that were clinically inappropriate.

**Table 1:** Participant Demographics, Background and Prior Experience with Artificial Intelligence.

| Characteristic | No. (%) |
| --- | --- |
| **Surgery Performed** |  |
| Laparoscopic sleeve gastrectomy | 12 (75%) |
| Laparoscopic Roux-en-Y gastric bypass | 4 (25%) |
| **Sex** |  |
| Female | 13 (81%) |
| Male | 3 (19%) |
| **Race/Ethnicity** |  |
| Hispanic | 9 (56%) |
| Non-Hispanic White | 4 (25%) |
| Non-Hispanic Black | 3 (19%) |
| **Age Group, years** |  |
| 18–24 | 1 (6%) |
| 25–34 | 4 (25%) |
| 35–44 | 4 (25%) |
| 45–54 | 0 (0%) |
| 55–64 | 5 (31%) |
| ≥65 | 2 (12%) |
| **Highest Education Level Completed** |  |
| High school graduate or equivalent | 3 (19%) |
| Some college | 3 (19%) |
| College degree | 6 (38%) |
| Advanced graduate degree | 4 (25%) |
| **Total Annual Household Income** |  |
| \$20,001–\$50,000 | 3 (19%) |
| \$50,001–\$100,000 | 5 (31%) |
| \$100,001–\$200,000 | 1 (6%) |
| >\$200,000 | 2 (12%) |
| Prefer not to answer | 5 (31%) |
| **Previously Used Information Sources** |  |
| Google | 16 (100%) |
| Physician | 9 (56%) |
| Friends | 4 (25%) |
| YouTube | 3 (19%) |
| TikTok | 2 (12%) |
| Support Groups | 2 (12%) |
| Hospital Websites | 2 (12%) |
| Dietitian | 1 (6%) |
| Family | 1 (6%) |
| Pamphlets | 1 (6%) |
| **Prior Experience with LLMs** |  |
| No Experience | 9 (56%) |
| A Little Bit of Experience | 5 (31%) |
| Some Experience | 1 (6%) |
| Quite a Bit of Experience | 1 (6%) |
| A Lot of Experience | 0 (0%) |
| **LLM Usage in the past 12 months** |  |
| Did Not Use | 9 (56%) |
| A Few Times | 4 (25%) |
| Once a Month | 0 (0%) |
| 1-2 times a week | 2 (12%) |
| Daily | 1 (6%) |

LLMs: Large Language Models.

### Qualitative Findings

We identified four overarching themes from interviews with adult patients undergoing bariatric surgery on their perceptions of BariBot usability following surgery: accessibility and usability, quality and consistency of information, perceived trust, and anticipated future use. Through interviews, participants contrasted BariBot with commonly used information sources, including search engines, websites, social media, and clinical encounters.

### Accessibility and Usability

#### Efficient Access to Structured Information

Participants described BariBot overall as accessible, efficient, and easy to use. In contrast to prior experiences with online information platforms, which were often characterized as fragmented and time-consuming, BariBot was perceived as providing direct responses to participant inquiries with minimal effort when prompted. The immediacy of BariBot’s responses was cited as a key strength, with several noting “Timewise, it saved me a ton of time” to look up and sort through information online, and “it was very fast and very simple.” (40–59-year-old participants). Participants appreciated being able to use the application at any hour, especially during recovery periods when clinical support from physicians and care teams may not be readily available: “What’s available right now at three in the morning? … I would have used it [BariBot].” (18–39-year-old participant) The interface was considered intuitive, and the use of bullet-point formatting was perceived as a feature that enhanced readability: “I think the way it’s responding back was good…short little statements…bullet points…it was concise and to the point where you don’t have to read too much but you got the understanding.” (40–59-year-old participant) A 40–59-year-old participant reported, “it gave me key information…based on the questions that I actually input.” Multiple others described the responses as “step by step,” “direct,” and “non-repetitive.”

### Conversational Flow and Contextual Follow-up

Some participants noted that the application often anticipated follow-up questions, further streamlining the experience. One 40–59-year-old participant stated, “at the end, [BariBot] provid[ed] additional information or questions …it automatically knew what I might be asking next.” A 40–59-year-old participant mentioned: “I just kept on going and going because it tells you itself - ‘Would you like something similar to this?’ … and it was very helpful…” Participants valued the application’s conversational flow and personalized tone, which made it feel more interactive than static web pages. Several compared it to “a computer-human,” and “talking to someone you know,” and appreciated its ability to carry context from one question to the next.

### Private and Judgment-Free Question Asking

Participants also described BariBot as reducing the social pressure sometimes associated with asking questions during clinical encounters. A 40–59-year-old participant reflected that “sometimes doctors get annoyed with maybe certain questions…nobody wants to be made to feel like they’re kind of an idiot,” and noted that BariBot “takes the anxiety [out] about asking certain questions.” Similarly, an 18–39-year-old participant described the application as “more…private,” adding, “I could be comfortable and not feel awkward asking the questions…I’m more of [an] introverted person so it’s easier for me to communicate like that instead of like talking to a person.” Participants therefore perceived BariBot as a private, judgment-free setting for postoperative questions.

### Suggested Improvements and Accessibility Needs

Participants suggested improvements to BariBot, including increasing the font size and tailoring information by surgery type for more personalized responses. An 18–39-year-old participant recommended adding “text to speech, if that’s an option” as a feature and recommended the application “..give more of like just context of where it’s from…we sourced this from reference”. Others also recommended providing source citations to enhance transparency. One 60-79 year-old participant reported Google was easier to navigate compared to BariBot due to a preference for “go[ing] through the different results as opposed to just being given the information [from a single source]” and recommended the application “introduce them[selves] or what it is” to make it more personable to patients.

### Quality and Consistency of Information

Participants described BariBot’s responses as clear, specific, and consistent with post-op information they had previously received from the clinical team. Several participants contrasted this experience with internet searches, which they perceived as fragmented or internally inconsistent. A 40–59-year-old participant explained that the information she found when searching online sources for postoperative bariatric surgery care “wasn’t correct. Sometimes it was over-exaggerated. And to me, sometimes it had false information.” Furthermore, an 18–39-year-old participant noted discrepancies across websites, stating, “different articles would say something different as far as what was the recovery, what are the side effects [of the surgery]? What are the benefits?” Similarly, another participant reported a lack of clarity when searching online, describing online content available as “vague. It wasn’t precise … nothing was uniform. A lot of it was just trying to compare [websites]… just to get one [answer]” (40–59-year-old participant).

In contrast, BariBot was perceived as providing organized and directly applicable information. An 18–39-year-old participant stated, “It was pretty accurate as far as stuff I’ve learned before.” Participants also emphasized that BariBot reduced the need to synthesize or cross-check information across multiple sources. One 40–59-year-old participant stated, “I don’t really feel like I have to cross-check the information like I would on Google,” while a 40–59-year-old participant noted that BariBot provided information that did not feel incomplete: “It gave me information that wouldn’t be considered missing…because you find a lot of incorrect information online.”

### Perceived Trust

#### Conditional Trust and Institutional Credibility

Participants described trust in BariBot as shaped by its consistency with prior clinical guidance and its association with a reputable clinical institution. One 18–39-year-old participant stated, “I’m continuing to open up to it. Especially this…coming from a reputable hospital too, and…them [hospital] being open to using it [BariBot].” A 40–59-year-old participant described BariBot’s responses as “aligned with everything else I was hearing from the professionals… between all of the packets I got from the nutritionist and all the different people who came in,” valuing “the reiteration,” rather than a new presentation of information. Several participants emphasized that this alignment made the application feel more reliable than general internet searches.

#### Educational Adjunct Rather Than Clinician Replacement

At the same time, participants distinguished between using BariBot for general postoperative education and relying on it for urgent or individualized medical decision-making. One 40–59-year-old participant stated, “If I’m having, like, an actual medical emergency, I’m not gonna mess with that,” underscoring that the application was viewed as a supplement rather than a replacement for clinician care. Participants also valued prompts that encouraged them to “not just rely on…the [BariBot] answers, but also consult with your doctor or surgeon” (18–39-year-old participant). Participants reiterated that the application should supplement, rather than replace, physician guidance. A 40–59-year-old participant stated “you still need to see your doctor. Nothing is gonna substitute that. But this is very helpful and to have just as a backup. But doctors are not replaceable…a doctor will see you and look at you and see what’s wrong with you or what you need. AI is not looking at you”. A 40–59-year-old participant shared “…so I think that it could like if it’s like a I need some more options and like diet or something like that, I could go on there like some easy questions like that.” Overall, participants viewed BariBot as a potentially useful adjunct for postoperative education, particularly as a source of accessible support between formal clinical encounters.

#### Transparency Concerns and Privacy Reassurance

Concerns about AI transparency remained common. A 60–79-year-old participant asked, “Where are they getting their information?” Another 40–59-year-old participant described AI as “a double-edged sword,” noting concern that the system had “no real moral compass” and depended on “the things that it’s been taught.” These concerns were partly offset by privacy-related reassurance. Several participants noted that BariBot did not request personal information; one 40–59-year-old participant stated that the application “doesn’t ask me any of my personal information,” which made the interaction feel less concerning. Participants described trusting BariBot more than broad internet searches but still described a desire for verification. An 18–39-year-old participant stated, “Maybe I trust it a little bit more than Google, but I still want to verify it.” Participants described trust in BariBot as often conditional rather than absolute, and they saw it as an improvement over general internet searches, which they felt still needed to be subjected to independent verification.

### Anticipated Future Use

Participants expressed interest in the continued use of BariBot beyond the inpatient postoperative period. Several described the application as a tool they would use at home if it were available. One 60–79-year-old participant stated, “I would take it home,” and an 18–39-year-old participant similarly noted, “If I was able to download it, I would have downloaded it already.” Participants also viewed BariBot as useful for questions that arise between scheduled clinical encounters. A 40–59-year-old participant described the application as “an in-between,” explaining that it could help if she needed “some more options on diet” or “a meal plan for the day.” Others described potential use for postoperative diet progression, symptom clarification, exercise, hydration, recipes, and calorie tracking. One 40–59-year-old participant extended this concept beyond bariatric surgery, stating, “I would probably see me using it after any office visit,” and cited a chronic orthopedic issue as another context in which an AI-based educational tool could be useful. One 40–59-year-old participant emphasized the importance of updated content, stating, “You want to see what’s current, what’s happening out there.”

## DISCUSSION

In this prospective, single-arm mixed-methods feasibility study, an AI-powered educational application for patients undergoing bariatric surgery was found feasible and associated with high perceived usability and acceptability. All participants completed the application use session and post-intervention assessments. No technical failures or patient-requested discontinuations occurred, and no clinically inappropriate responses were identified during physician review of patient-application interactions. Qualitative analysis of post-intervention interviews showed that participants perceived BariBot as accessible, efficient, private, and useful for obtaining structured postoperative information. Importantly, participants valued not only the content of individual responses, but also the application’s ability to support conversational continuity, follow-up questioning, contextual responses, and suggested related topics. Participants expressed cautious trust in the application, viewing it as more targeted than general internet searches while emphasizing the need for transparency, clinical oversight, and use as a supplement rather than a replacement for clinician guidance.

Our findings build on prior literature evaluating LLMs for patient education in bariatric surgery, most of which have relied on simulated questions, expert review, or benchmark-style assessments rather than direct patient use. (31,34,35,48–50) In contrast, this study evaluated patient-directed interaction with an LLM-powered educational application during the immediate postoperative period, when questions about diet, symptoms, medications, activity, and recovery commonly arise. By allowing participants to engage in extended, self-directed conversations with BariBot, we captured dimensions of the patient experience that are difficult to assess using isolated prompts or benchmark questions. This distinction is important because feasibility, usability, trust, and perceived value cannot be fully assessed through static prompts or clinician adjudication alone.

Participants valued features intrinsic to conversational systems, including contextual continuity, follow-up questioning, structured responses, and suggested related topics suggesting the value of LLM-based education may lie not only in the accuracy of individual responses, but also in the ability to support iterative, patient-centered information seeking between clinical encounters. Participants valued the application’s ability to preserve conversational context, respond to follow-up questions, and suggest relevant next topics for discussion, features that made the interaction feel more responsive and personalized than static educational resources or single-query information retrieval. Furthermore, the application’s language was adapted to individual patient education level, a strategy supported by our prior work showing unprompted LLM responses frequently exceed recommended text readability thresholds. (43,44,51) By tailoring language complexity and presenting information in a clear, structured format, BariBot appeared to reduce cognitive burden and promote engagement. The findings therefore provide preliminary evidence that an LLM-based application can be implemented in a real-world surgical workflow and can be perceived by patients as accessible, understandable, and useful.

Trust emerged as a central consideration in participants’ assessment of BariBot. Participants appeared more willing to engage with the application when its responses aligned with prior clinical guidance, when it was perceived as connected to a reputable health system, and when it did not request personal information, similar to prior research. (52,53) At the same time, participants expressed concern regarding the transparency of AI-generated information and distinguished educational support from urgent or individualized medical decision-making. These findings suggest that successful implementation will depend not only on response accuracy, but also on clear disclosure of the application’s role, explicit limits on its intended use, preservation of clinician oversight, and mechanisms that direct patients to the clinical team when questions involve potentially urgent or high-risk concerns.

The anticipated value of BariBot appeared greatest for questions arising between formal clinical encounters, particularly those related to diet progression, hydration, medications, physical activity, symptom interpretation, and postoperative expectations. These topics often require repeated reinforcement and may generate uncertainty after discharge, when immediate access to the clinical team is limited. However, implementation should not simply add an AI application to existing workflows without considering how it interfaces with established educational materials, patient messaging systems, dietitian support, and postoperative triage pathways. Future improvements should provide procedure-specific guidance, incorporate source citations, support accessibility features such as larger font sizes and text-to-speech functionality, and remain aligned with institutionally approved recommendations.

While physician review of participant-BariBot interactions identified no clinically inappropriate or potentially harmful responses, this finding should be interpreted cautiously. The present study evaluated a limited number of interactions during a brief period of supervised inpatient use and was not designed to establish the safety of unrestricted or longitudinal use after hospital discharge. LLM-generated responses may vary according to the wording of a question, the preceding conversational context, model updates, and the complexity or urgency of the clinical scenario. In routine postoperative care, patients may present ambiguous symptoms, omit clinically important details, or seek reassurance regarding evolving complications, creating a risk that an educational application could provide incomplete guidance or delay appropriate evaluation. Future implementation should therefore incorporate explicit escalation pathways, clear instructions regarding symptoms that require urgent clinical attention, ongoing review of application interactions, and formal governance processes to monitor performance after model or content updates. The recent, rapidly evolving agentic application architectures may facilitate these safeguards by assigning specialized components to tasks such as retrieval of institutionally approved information, identification of potentially high-risk symptoms, verification of generated responses, and escalation to the clinical team when predefined risk thresholds are met. (54,55) However, such systems may also introduce new sources of error and should undergo prospective validation before being relied upon for autonomous quality control or clinical escalation.

Because BariBot was evaluated only among English-speaking participants using a text-based interface, its usability, acceptability, and safety cannot be assumed across languages or among patients with visual, cognitive, motor, or digital-access limitations. This consideration is particularly important because LLM performance is not language invariant; prior studies have identified differences in the accuracy, comprehensibility, and reliability of medical information generated across English and non-English languages. (56–58) Future versions should therefore undergo language-specific evaluation rather than relying solely on direct translation of English-language content. Such efforts will be necessary to ensure that patient-facing AI applications expand access to reliable education rather than reinforce existing disparities in health information access.

This study has several limitations. First, it was conducted at a single tertiary academic center with a small sample size, which may limit the generalizability of the findings to broader bariatric populations and other care settings. Although the sample was small given the design as a feasibility study, participants represented varied age groups, racial and ethnic backgrounds, educational attainment, and prior experience with LLMs, supporting the preliminary applicability of the findings across patients with differing demographic and technological backgrounds. Second, the study included only English-speaking participants who were able to engage with a text-based digital interface, excluding individuals who spoke languages other than English, had visual impairments, or faced other barriers to technology use. Third, the brief inpatient evaluation precluded assessment of long-term use, engagement, knowledge retention, adherence, and clinical outcomes. Finally, the study was designed as a feasibility assessment and lacked a comparator group, which limits inference regarding whether the application performed better than standard educational resources alone.

## CONCLUSION

BariBot was feasible to implement and was associated with high perceived usability and acceptability among patients undergoing bariatric surgery. Participants valued its accessibility, conversational responsiveness, privacy, and consistency with prior clinical guidance, while expressing cautious trust and a continued preference for clinician involvement in urgent or individualized medical decisions. These findings support further investigations into the development of AI-based educational applications as adjuncts to postoperative care. Larger, longitudinal, and comparative studies are needed to determine whether such applications can be implemented safely and whether they improve patient understanding, engagement, or clinical outcomes.

## Data Availability

The data generated and analyzed in this study are not publicly available because they contain sensitive participant information, including qualitative interview and application-interaction data, and their disclosure could compromise participant privacy and confidentiality. Aggregate findings relevant to the study are presented in the manuscript.

## Author Contributions

J.S.S.: Co-conceptualization, co-designed the analyses, performed statistical analysis, co-development of application, co-wrote the manuscript, and approved the final draft.

S.N.: Co-conceptualization, co-designed the analyses, performed data collection, performed statistical analysis, co-wrote the manuscript, and approved the final draft.

S.H.: Co-designed the analyses, performed statistical analysis, edited the manuscript for important intellectual content, and approved the final draft.

A.T.: Co-conceptualization, co-designed the analyses, co-development of application, performed data collection, edited the manuscript for important intellectual content and approved the final draft.

R.C.C.: Supervision of qualitative research design and analysis, edited the manuscript for important intellectual content, and approved the final draft.

N.W.: Supervision of qualitative research design and analysis, edited the manuscript for important intellectual content, and approved the final draft.

T.Y.: Performed data collection, edited the manuscript for important intellectual content and approved the final draft.

D.C.: Performed data collection, edited the manuscript for important intellectual content and approved the final draft.

N.R.: Performed data collection, edited the manuscript for important intellectual content and approved the final draft.

R.A.: Edited the manuscript for important intellectual content and approved the final draft.

R.W.: Edited the manuscript for important intellectual content and approved the final draft.

B.A.: Edited the manuscript for important intellectual content and approved the final draft.

K.S.: Edited the manuscript for important intellectual content and approved the final draft.

H.W.: Edited the manuscript for important intellectual content and approved the final draft.

N.P.T.: Edited the manuscript for important intellectual content and approved the final draft.

K.S.: Supervision, co-conceptualization, co-designed the analyses, edited the manuscript for important intellectual content and approved the final draft.

## AI writing statement

During the preparation of this work, the authors used various GPT models to enhance readability and language. After using these tools/services, the authors reviewed and edited the content and take full responsibility for the content of the publication.

## Abbreviations

AI: Artificial Intelligence
LLM: Large Language Model
MBS: Metabolic and Bariatric Surgery
POD1: Postoperative Day 1
SUS: System Usability Scale

