## Supplementary Appendix for "Evaluation of an AI-Powered Patient Education Application in Bariatric Surgery: A Prospective Mixed-Methods Feasibility Study"

**Appendix 1: Pre-Intervention Survey**

What is your age in years? ___________

What is your gender identity? (Check all that apply)

□ Female – Cisgender (identifying as gender assigned at birth)

□ Male – Cisgender (identifying as gender assigned at birth)

□ Female – Transgender

□ Male – Transgender

□ Non-binary/third gender

□ Prefer to self-describe (please specify): ________________________

□ Prefer not to say

Are you of Hispanic, Latino, or Spanish origin?

o Yes

o No

What is your race?

o American Indian or Alaska Native

o Asian

o Black or African American

o Native Hawaiian or Other Pacific Islander

o White

o Other

What is the highest level of education you have completed?

o 8th grade or less

o Some high school

o High school graduate or equivalent

o Some college

o College degree

o Advanced graduate degree

What is your best estimate of your household’s total annual income?

1) Less than $10,000

2) Between $10,000 to $20,000

3) Between $20,001 to $50,000

4) Between $50,001 to $100,000

5) Between $100,001 to $200,000

6) More than $200,000

7) Prefer not to answer

Through which of the following sources have you been primarily obtaining

information related to obesity and the surgery itself? Please select all that

apply:

o Healthcare professionals (e.g., doctors, nurses)

o Official medical or health websites (e.g., WebMD, Mayo Clinic)

o Social media platforms (e.g., Facebook, Instagram, Twitter)

o Online forums or support groups (e.g., Reddit, specialized support groups)

o Friends or family members who have undergone similar procedures

o Printed materials (e.g., brochures, books, magazines)

o Educational seminars or workshops

o Other (please specify)

Which of the following best describes your level of experience using artificial

intelligence large language models (Example ChatGPT, BARD, Gemini, Claude) ?

o No experience

o A little bit of experience

o Some experience

o Quite a bit of experience

o A lot of experience

Approximately how often did you use a large language model (Example

ChatGPT, BARD, Gemini, Claude) in the past 12 months?

o Daily

o 1-2 times a week

o Once a month

o A few times

o Did not use ChatGPT in the past 12 months

In the past 12 months, what have you used a large language model (Example ChatGPT, BARD, Gemini, Claude) for?

**Appendix 2: Interview Guide**

When you think about searching for information related to managing your weight or the surgery itself prior to today in general, what comes to mind?

- *How would you describe your experience finding information related to obesity and bariatric surgery before today?*
  - *How would you decide what you could trust vs not?*
- *What sources of information did you use prior to your surgery?*
  - *Overall, how was your experience with these sources?*
  - *How is the quality of the information you find? How do you judge the quality?*
  - *How are they the same? Is it different in any way?*
  - *Did you have any particular likes or dislikes about searching for information or the sources you used?*
- *How would you describe the difficulty/ease in finding information?*
  - *time required to find it?*
  - *ease of understanding of the materials?*

Think back to your views about large language models like ChatGPT, BARD or Gemini-pro in general.

- How familiar are you with AI?
- How would you compare your prior views to your experience today?
- How did your experience today change your views on AI?

What was your experience with using the artificial intelligence large language model today?

Overall, how was your experience with this program?

- *Would you recommend any changes to the program?*
  - *What specific aspects did you find most helpful?*
  - *Were there any aspects you found that were lacking?*
  - *How easy is it to use the application compared to other sources?*
    - *How easy was it to understand the information?*
    - *How did you feel about the amount of information presented? The organization?*
  - *Compared to other sources, do you feel this is a different personalization or experience? How is the dynamic/interaction different?*
- *Can you see yourself using this type of technology in the future for your healthcare? Tell us why*
  - *What would you use it for?*
  - *Are there any barriers that would prevent you from using AI in the future?*
- *Would you trust validated and tested artificial intelligence sources for healthcare information in the future?*
  - *How much do you trust AI versus Google versus family/other sources*
  - *Is validation important to you? By who?*
  - *What would it take for you to feel validated?*
- *Were there any additional recommendations you would give to the improve artificial intelligence that you used today?*
  - *What features would you add to make it more useful*
  - *Are there any ways it could be more personalized?*
  - *Could your interaction with the model be improved in any way?*
  - *Anything that would make you want to use it more? Incorporate it as a regular source of information?*
- *Do you have any concerns about this technology?*
  - *Any privacy concerns?*
- Is there anything else you think we should be asking you?
- Is there anything else you want to say?
- Would it be okay if we used your responses in this interview/with the chatbot in a published paper (identities concealed)?
